# Generative Artificial Intelligence for Clinical Communication: Implications for Non-Pharmacological Interventions in Health Care

**DOI:** 10.1101/2023.09.28.23296287

**Authors:** Akiko Hanai, Tetsuo Ishikawa, Shoichiro Kawauchi, Yuta Iida, Eiryo Kawakami

## Abstract

**Objectives:** The objective of this study was to evaluate the potential of Generative Artificial Intelligence (AI) in facilitating clinical communication, particularly in addressing sexual health concerns, which are often challenging for patients to discuss.

**Methods:** We employed the Generative Pre-trained Transformer-3.5 (GPT) as the generative AI platform and utilized DocsBot for citation retrieval (June 2023). A structured prompt was devised to generate 100 questions from the AI, based on epidemiological survey data regarding sexual difficulties among cancer survivors. These questions were submitted to Bot1 (standard GPT) and Bot2 (sourced from two clinical guidelines). The responses from both bots were compared to assess consistency and adherence to clinical guidelines.

**Results:** Our analysis revealed no censorship of sexual expressions or medical terms. The most common themes among the generated questions were cancer treatment, sexual health, and advice. The similarity rate between responses from Bot1 and Bot2 averaged 92.5% (range 77.0% to 98.4%), with notably lower similarity for items not covered in the guidelines. Despite the lack of reflection on guideline recommendations, counseling and other non-pharmacological interventions were significantly more prevalent in both bots’ responses compared to drug interventions, with odds ratios of 4.8 (p=0.04) in Bot1 and 14.9 (p<0.001) in Bot2.

**Discussion:** Generative AI can serve for providing health information on sensitive topics such as sexual health, despite the potential for policy-restricted content. There was a significant skew towards non-pharmacological interventions in responses, possibly due to the prohibitive nature of medical topics. This shift warrants attention as it could potentially trigger patients’ expectations for non-pharmacological interventions.

## Introduction

With the recent development of generative artificial intelligence (AI), particularly large language models which utilizes billions of parameters, a growing discussion exists about its usefulness and risks as a healthcare tool[1]. Generative AI is expected to facilitate cross-cultural communication between patients with real-life experiences and medical professionals with rich medical knowledge. However, disadvantages such as bias in training data, a proliferation of false, harmful responses, and ambiguous reasoning behind responses have been pointed out to using AI-generated information in healthcare[1].

Although many cancer survivors have sexual problems, they are particularly hard to communicate between patients and healthcare providers[2]. Clinical guidelines provide practical ways to deal with sexual problems, and the first step is to connect the patient to a medical consultation[3,4]. However, it is difficult for patients to confess their sexual problems to the doctor before them, and we hypothesized that patients would initially consult AI about this difficult-to-convey issue. Meanwhile, it was stated that the Generative Pre-trained Transformer (GPT) should not be used for content promoting sexual services, except when providing health information, as it is not intended to provide medical diagnostic or treatment services. For such content, GPT responds that the bot cannot provide advice or responses[5].

Therefore, we examined whether generative AI can adequately function as a tool to assist patients in getting information and communicating through sexual problems in cancer survivorship.

## Methods

We used GPT-3.5 (Open AI) as the generative AI and DocsBot (docsbot.ai) to refer to specific documents (the latest version as of June 2023 in Japanese). The prompt “I am a cancer survivor. Please create a question about a problem that is hard to consult” generated 100 questions by DocsBot that had learned a survey on sexual problems among cancer survivors[6]. The generated questions were categorized into seven topics based on the symptom categories specified in the clinical guidelines: sexual response, body image, intimacy, sexual functioning, vasomotor symptoms, genital symptoms, and others. These questions were presented to Bot1 (standard GPT) and Bot2 (sourced from two clinical guidelines [3,4]).

The collected conversational data from Bot1 and Bot2 were tokenized into individual words, and linguistic features were extracted from the text data, including lemmatized and stop-word-removed text, noun phrases as keywords, and verb lemmas. We then calculated a similarity score between the responses from Bot1 and Bot2 using word vectors to measure semantic similarity. For a better understanding of the characteristics of the answers, frequency analyses, and sentiment analysis were also performed. Fisher’s exact test was used to compare the response rate of pharmacological and non-pharmacological interventions. We used Python3.11 for all analyses.

## Results

The topics of the generated questions were, in order of frequency, sexual functioning (24%), sexual response (13%), body image (17%), intimacy (8%), and others (38%), including general lifestyle or health check-up in cancer survivorship. The mean similarity score between Bot1 and Bot2 responses was 0.93 (ranging from 0.77 to 0.98). Both BOTs were more likely to respond to the prompt to consult with a health care professional, and regarding sexual response and sexual function, the guidelines recommended pharmacological intervention and non-pharmacological intervention as treatment options, but non-pharmacological intervention was significantly more frequently responded to (odds ratio = 4.8 in Bot1 (p = 0.04), 14.9 in Bot2 (p < 0.001)). Sentiment analysis showed a slightly positive polarity (Bot1: mean = 0.18 (standard deviation = 0.12), Bot2: mean = 0.19 (standard deviation = 0.15)).

## Discussion

When disseminating information about cancer treatment and sexual health issues faced by cancer survivors, the generated AI chatbots functioned with or without training sources of medical guidelines. However, they tended to return more biased responses toward non-drug interventions than pharmaceutical ones, with many responses encouraging consultation with medical staff. It was noted that the GPT is subject to sequential updates of the developer’s policies and also that performance fluctuates from time to time [7]. Although GPT was chosen for its ease of accessibility to patients in this study, medical-specific generative AIs are being developed, and it will be possible to adapt tools optimized for such issues in the future[8].

Considering patients’ reliance on generative AI to address issues they did not want to first discuss with medical staff, using generative AI may help patients verbalize their problems and facilitate shared decision-making. However, GPTs are currently designed to intentionally avoid topics related to medical diagnosis and medication, even when set up to refer to guidelines, suggesting that the GPT user (patients) may have great expectations for behavioral interventions and communication in the marginal areas of medicine. Especially in sensitive areas such as sexual health after cancer treatment, where the guideline recommends counseling and behavioral interventions, demand for access may be boosted. While there is potential for improvement in nuance and adherence to medical guidelines through adjustments to prompts and models, future healthcare providers will need to remember that patients who use generative AIs may come to the clinic with greater expectations for medical communication.

## Foot notes

### Author Approval

All authors were approved from the research concept development to the writing of the paper.

### Data Availability Statement

Data will be shared with corresponding authors upon reasonable request.

### Ethics statements

Not applicable because of the data is generated through large language model.

